# A New Approach to the Dynamic Modeling of an Infectious Disease

**DOI:** 10.1101/2020.10.30.20223305

**Authors:** B Shayak, Mohit M Sharma

## Abstract

In this work we propose a delay differential equation as a lumped parameter or compartmental infectious disease model featuring high descriptive and predictive capability, extremely high adaptability and low computational requirement. Whereas the model has been developed in the context of COVID-19, it is general enough to be applicable mutatis mutandis to other diseases as well. Our fundamental modeling philosophy consists of a decoupling of public health intervention effects, immune response effects and intrinsic infection properties into separate terms. All parameters in the model are directly related to the disease and its management; we can measure or calculate their values a priori basis our knowledge of the phenomena involved, instead of having to extrapolate them from solution curves. Our model can accurately predict the effects of applying or withdrawing interventions, individually or in combination, and can quickly accommodate any newly released information regarding, for example, the infection properties and the immune response to an emerging infectious disease. After demonstrating that the baseline model can successfully explain the COVID-19 case trajectories observed all over the world, we systematically show how the model can be expanded to account for heterogeneous transmissibility, detailed contact tracing drives, mass testing endeavours and immune responses featuring different combinations of limited-time sterilizing immunity, severity-reducing immunity and antibody dependent enhancement.

## INTRODUCTION

We split this discussion into two Sections, one general and the second more specific.

### §1 OVERVIEW OF DISEASE MODELING APPROACHES

With the spread of COVID-19 like wildfire all over the world, infectious disease dynamics has suddenly been promoted from a niche area of dynamical systems theory to the foremost topic in applied mathematics and sciences research. Mathematical modeling is the only scientific tool which allows us to predict the trajectories of the disease in advance and take intervention measures accordingly. There are four approaches to modeling of an infectious disease, which we describe in summary below.

- **Lumped parameter or compartmental model:** These are differential equation models, like S-I-R and S-E-I-R. In most cases these models use ordinary differential equations (ODE), some though not many use delay differential equations (DDE) and a few use partial differential equations (PDE). These models are deterministic in that the equations do not involve random variables. The advantage of these models is that they are physically insightful and computationally tractable (especially ODE and DDE models). The drawback is that the division of population into compartments automatically requires averagings and assumptions; the extent to which these limitations fetter the performance of the model depends to a large degree on the model itself. We shall elaborate on this type of model in the next Section.
- **Agent-based model:** This model considers people as lattice sites on a network. Each site can be in one of several states – typical states are healthy and susceptible, exposed and non-infectious, asymptomatic infectious and at large, symptomatic and quarantined etc. A lattice site contracts the infection with a certain user-defined probability if its one or more neighbours are infectious, and then progresses through the successive states with user-defined probabilities and durations. The advantage of this model is that it is the closest representation of reality and hence is capable of extreme accuracy. Thus for example, it can incorporate a sophisticated contact tracing effort with multi-level two-way tracing, or predict the effects of a single unlicensed party. The disadvantages are enormous computational cost, lack of physical insight into the results and sensitivity of the results to the underlying network structures assumed by the modellers. Among prominent examples of agent-based models are the studies conducted by the London School of Hygiene and Tropical Medicine [1], Imperial College [2] and Los Alamos National Laboratory [3]. Another such model [4] has been able to explain the linear growth in corona cases seen in many regions of the world – linear disease trajectory is not seen in differential equation models except as a marginal case.
- **Stochastic differential equation model:** These attempt to combine the features of lumped parameter and agent based models, by writing differential equations which feature random variables. Examples are the Cornell University model [5] and the Jadavpur University model [6]. Our personal preference is for the preceding two kinds of models since they are more direct and intuitive.
- **Data-driven model:** These models simply take the existing data of COVID-19 spread over the past week or month (or longer) and use machine learning etc methods to generate a forecast for the next week or month. They pay little or no attention to the underlying processes driving the spread of the disease. The ease of preparing these models for (at least potential) publication contributes to their popularity and their huge numbers in the literature; at least one of these efforts [7] has failed on the long term to live up to the hype which it initially generated.

This concludes our summary of the different approaches in existence to the mathematical modeling of infectious diseases in general and COVID-19 in particular. What we propose in this Article is a lumped parameter model; we shall now present a summary of the state of the art in this area.

### §2 LUMPED PARAMETER MODELS

The first ever model for an infectious disease was of this type – it was the S-I-R model invented in 1927 by WILLIAM KERMACK and ALEXANDER MCKENDRICK [8] for generating the epidemiological curves of plague. A hundred years later, this model has been applied directly to COVID-19 as well. More common for this disease however is the S-E-I-R model, which we describe briefly. The four compartments here stand for susceptible, exposed, infected and removed (recovered or dead), and the equations are

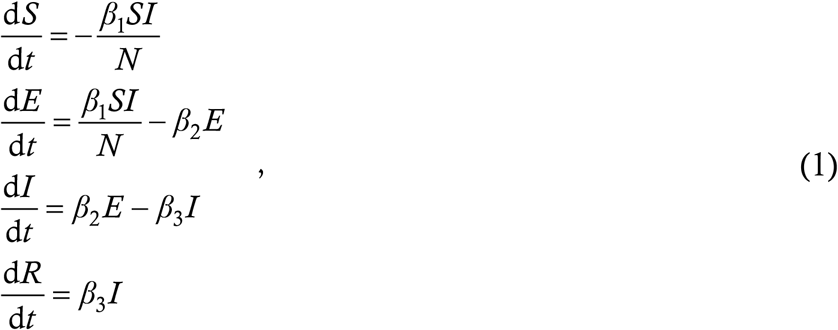

where the *β*’s are constants. Although all the *β*’s look like rates, they actually represent two kinds of real-world phenomenon. *β*_1_ for example is a true rate; it is the number of people whom one infected person infects in one unit of time. *β*_3_ however is not a true rate but a reciprocal time – the fourth equation d*R*/d*t* = *β*_3_*I* attempts to represent the fact that patients remain infected for an average duration of 1/*β*_3_ before being removed from consideration. This is a conceptual stretch; for if at *t* = 0 we have a certain number *I*_0_ of infected people and set *β*_2_ = 0 (by whatever means), then the subsequent infection profile looks like *I* = *I*_0_ exp(−*β*_3_*t*). In reality however, if there are *I*_0_ infected people today and no fresh infections, then they will all recover more or less uniformly within the next 10 or 14 days, or whatever is the infection period – there will not be a wave of recoveries at the start and a small but finite number of infected people several months down the line. This is a limitation of lumped parameter ODE models.

A second limitation of these models is that the parameters are often heuristic and not related directly to real-world phenomena governing the disease. For example, you may ask the question, “Suppose it turns out that the asymptomatic infection period, which was earlier thought to be 4 days, is actually 7 days, then which *β*’s would have to change and by how much ?” or equivalently “Suppose the authorities of a university campus were to initiate mandatory weekly testing of all students, then which *β*’s would change and by how much ?” These questions are all but impossible to answer for the basic S-E-I-R model (1).

More elaborate models with eight or more dependent variables and even more parameters [9,10] have been set up in attempts to answer such questions – in such cases, the conceptual clarity of the model and the results arising therefrom potentially take a hit. For example, a recent study featuring such a model [11], led by an extremely distinguished mathematician, has been challenged [12] on the grounds that the parameters in the model “usually are not constrained in any [way] by our understanding of COVID-19 epidemiology” and that “a few of them are varied throughout a wide range, from 67 to 4,75,000 in [one instance, with] no justification being given for this”. Whereas it is not our aim to critique either the original study or the rebuttal, we do believe that a model whose basic parameters are concrete things like asymptomatic fraction and latent infectious period is immune to attacks of this nature. Moreover, an ODE model with excessively many parameters can generate good fits to existing case trajectories at many distinct points or regions of parameter space – future trajectories with the different parameter sets can vary widely however. The authors of one study [13] who had made a rash prediction on such a flimsy basis were taught their error the hard way [14].

DDEs have been used in the Literature much more sparingly than ODEs. A notable example [15] dates from the pre-COVID era (which already appears like some sort of distant dream); this model has been followed up and applied to COVID by a different set of authors [16]. Although the calculations in this latter paper are formidable, the conclusions – namely that one should always observe physical separation minima and wear a mask – are very mainstream. An elegant analytical solution to a linear DDE has been given in Ref. [17], which does not focus too much on the modeling aspects.

With this somewhat lengthy Introduction, we hope that we have been able to set the context for the present Article. We shall propose here a lumped parameter model for the spread of an infectious disease (not necessarily just corona) which has an elegant structure, is not over-ripe with parameters but at the same time directly incorporates the effects of realistic phenomena associated with the spread, such as latent transmission, asymptomatic carriers and test-trace-isolate programs. Our model further has the scope to accommodate different kinds of immune response to the disease such as permanent immunity, temporary sterilizing immunity, long-lasting severity-reducing immunity, antibody dependent enhancement etc. In Part 1 we present the derivation and solutions of the baseline model, in Part 2 we consider enhancements taking into account public health effects and in Part 3 we consider enhancements taking into account immunity effects. While much of the discussion has COVID-19 at its core, the features we model are quite general and are applicable to various infectious diseases such as plague, influenza, Ebola and anything else which the bats, cats and rats of the world may have in store for us in the future.

## 1 THE BASELINE MODEL

The treatment here closely follows our original works [18-20] in which we presented this model for the first time. We do not claim this Section as a novelty of the present Article, although we do believe that it is essential for what follows.

### §3 DERIVATION

We present the derivation in more detail than just a summary because it will form the building block of all the advanced variants we shall present in Parts 2 and 3. We focus on a region with good mixing among its inhabitants, such as a neighbourhood, town, village or city (it might be advantageous to break up a big city into several regions though, depending on connectivity). Our model features only a single dependent variable *y* (*t*) which is the cumulative count of corona cases in the region as a function of time. We measure time in days throughout this Article. Cases can be of two types – quarantined and at large. The former by definition have zero contribution to spread. The latter disseminate the virus freely to healthy and susceptible people via interaction. By interaction we mean both in-person interaction and interaction via objects (for example, a case contaminating an item at a grocery store which a subsequent healthy customer buys qualifies as an interaction). In what follows, we shall use the word **target** to denote a person with whom an at large case interacts. This spreading via interaction can be represented through the word equation

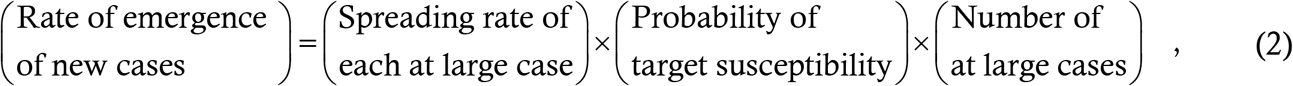

which we shall now express in a mathematical form.

Every at large case has a certain rate of interaction with targets. This interaction rate can vary widely – for example a grocer or banker might deal with 15-20 customers a day while a working from home software engineer might not interact with anyone at all in a week. For a lumped parameter model, we must average over various kinds of people; let the average interaction rate over all at large cases be *q*_0_ persons/day. This *q*_0_ depends on the degree of mobility in society – in a lockdown it will be far lower than in an unlocked state. Not every interaction however results in a transmission – for example a target sitting in the same classroom as a case might not contract the disease if both are wearing masks and sitting far enough apart. Similarly, a contaminated grocery item will not spread the virus to the customer if s/he sanitizes everything s/he buys. Let *P*_0_ be the probability (averaged over cases and targets) that an interaction between a case and a susceptible target actually results in a transmission. Note again the definition – **given that the target is susceptible**, *P*_0_ denotes the probability that s/he contracts the virus from the case. Then, we define the per-case spreading rate *m*_0_ = *q*_0_*P*_0_. We can see that *m*_0_ incorporates the effects of public health interventions.

Since *P*_0_ is conditional on the target’s being susceptible, we now need to account for the probability that s/he is not. The second term on the RHS of (2) factors this in. For the baseline model we assume permanent immunity i.e. we assume that recovered cases are insusceptible to further infection for all time. With permanent immunity, the probability of a random person’s being insusceptible is the total number of recovered cases divided by the total number of people in the region. The former is approximately *y* – it is actually a little less than *y* since not all cases at any time are recovered. But if the recovery duration is significantly less than the overall progression of the disease, then we can make the **instantaneous recovery** assumption and treat the recovery count as *y*. Let the total number of susceptible people in the region at the start of the epidemic be *N*. Then the probability of a random person’s being insusceptible is *y*/*N* and the probability of his/her being susceptible is 1 − *y*/*N*. The instantaneous recovery assumption ensures that our model possesses the very important property of d*y*/d*t* being identically zero when *y* = *N*, without having to incorporate a lot of fancy end-effects.

To motivate the third term, suppose hypothetically that all cases take a time *τ* days to recover and remain at large the whole time. Then the number of at large active cases now is exactly the number of people who fell sick between now and *τ* days back – mathematically, this is expressed as *y* (*t*) − *y* (*t*−*τ*). Here is the delay in the equation – note that we are using **delay** to express the recovery duration rather than an inverse rate, as in the S-E-I-R and related models. Of course, every case having the same recovery time and remaining at large throughout is simplistic. For one, symptomatic cases (with a few negligent exceptions) will generally go into quarantine after manifesting symptoms. For another, contact tracing drive will yield and isolate asymptomatic as well as pre-symptomatic or latent cases. We now partion the cases into three classes : (*a*) contact traced cases, (*b*) untraced symptomatic cases and (*c*) untraced asymptomatic cases. Let the number *μ*_1_ between 0 and 1 denote the fraction of total cases who are asymptomatic, let *μ*_3_ between 0 and 1 denote the fraction of total cases who do not get contact traced, let *τ*_1_ denote the asymptomatic infection period and *τ*_2_ the latency or pre-symptomaticity period during which a to-be-symptomatic case is transmissible prior to developing symptoms.

The class (*a*) or contact traced cases account for 1−*μ*_3_ of the total. This is a quasi-heuristic rather than a phenomena-driven parameter which suffices for the baseline model. For this model, we also make two assumptions whose effects counteract each other – the first is the assumption of **zero non-transmissible incubation period (NTI)** and the second is that of **instantaneous contact tracing**. The first assumption implies that if a target contracts the virus, then s/he begins transmitting immediately following exposure. The second assumption implies that public health authorities track down a person’s contacts as soon as they begin the tracing process. We have used these assumptions so as not to clutter the baseline model with parameters – in Part 2 we shall show how to relax them. If the contact tracing starts from freshly reporting symptomatic cases, then, with the assumptions in place and taking for granted that all cases transmit continuously and uniformly, the average duration that the secondary cases of the reporting cases remain at large is *τ*_2_/2. Class (*b*) or untraced symptomatic cases account for fraction *μ*_3_ (1−*μ*_1_) of the total cases and they remain at large for the latency period *τ*_2_ before manifesting symptoms and going into quarantine (the baseline model does not account for the actions of negligent individuals who we hope are few in number). Finally, class (*c*) or untraced asymptomatic cases account for fraction *μ*_3_*μ*_1_ of the total cases and these remain at large for the asymptomatic infection period *τ*_1_. Applying the *y*(*t*) − *y*(*t* − *τ*) argument to each class yields the mathematical representation of the third term on the RHS of (2) as

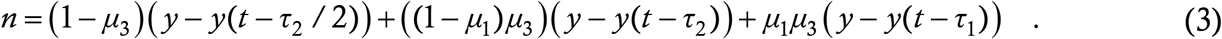

Multiplying all the terms on the RHS of (2) and simplifying the algebra in (3), we get

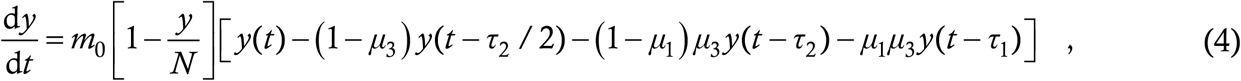

which is the **retarded logistic equation**, and the baseline model.

Equation (4) is a six-parameter model – the parameters are *m*_0_, *N, μ*_1_, *μ*_3_, *τ*_1_ and *τ*_2_. Among these, the last two are well-known – they are about 7 days and 3 days respectively [21]. **These are values we shall use throughout the entirety of this Article**. *N* is partially known since it is expected to be a sizeable fraction (say about 50 percent) of the region’s total population, especially if the region is relatively small and well-connected. The dependence of the trajectories on *N* is not very sensitive anyway. The asymptomatic fraction *μ*_1_ is partially-to-well known since each region has its own estimate of this parameter; this estimate is accurate or too low depending on whether contact tracing is extensive or poor and tests are abundant or scarce. *μ*_3_ is known to public health authorities who are aware of how many symptomatic cases they have detected through contact tracing and how many from walk-in tests – it is inaccessible to scientists who are not in contact with such authorities. Finally, *m*_0_ is difficult to measure a priori – although it can be estimated to some degree from mobility studies and/or surveys, it is more likely that given all the other parameters, it will have to be extrapolated from a data fit. Thus, an insider (i.e. a person with access to detailed information) basically has just one parameter to tune when attempting to fit a given data curve; an outsider has two or three. We can reasonably say that our model is frugal when it comes to parameters.

### §4 SOLUTIONS

We first note that (4) enables a direct calculation of the reproduction number *R* at any stage of evolution of the disease; the formula is

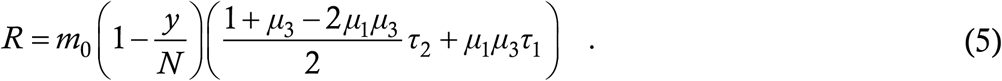

This follows from an elementary stability analysis which has been derived in the prior work [20,22] – the calculation of *R* from an ODE model [23] is non-trivial, and is in general limited to the starting value *R*_0_ only. We briefly present six classes of solutions of (4). These are also taken from prior work [18-20] and they do not need detailed explanations. We use numerical integration to solve (4), the method being second order Runge Kutta with a step size of 0·001 day. **We solve (4) in a Notional City having an initial susceptible population of *N* = 3**,**00**,**000 and an initial condition of zero cases to start with and 100 cases/day for the first seven days; such a City will be our solution domain in the entire Article**. For all Cities in this Section, we assume *μ*_1_ = 0·8. The parameter values of the six Cities A to F as well as real-world examples of each city are given in the Table below, while the solutions for these Cities are shown in the Figure which follows.

In each plot of the below Figure, we show three things – the case count *y*(*t*) as a blue line, its derivative 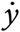(*t*) as a green line and the weekly case growth scaled down by a factor of 7 in grey bars. This latter is called the epidemiological curve or “epi-curve” and is of great interest to epidemiologists.

This shows us that the baseline model is itself capable of generating a diverse range of epidemic trajectories which we see in the real world. We now let the prior work be – the baseline model has been derived in a manner which will motivate the various extensions, and enough evidence has been accumulated that this model is realistic and has mathematical advantages relative to conventional lumped parameter models. Henceforth we focus on the extension of the baseline model to various scenarios which can and do arise in reality, in terms of both public health interventions and immune response.

## 2 PUBLIC HEALTH INTERVENTION EFFECTS

Here we build upon the baseline model to accommodate the effects of various public health interventions. Our focus shall be on the generalization process of the model itself, rather than on the solutions of these generalized models or the fitting of these solutions to different regions. This is because the augmented models will often be multiparameter and complex. As private researchers, we do not have access to accurate information regarding the parameter values, and for different combinations of these values, the solutions can show a wide variety of behaviour as well as non-unique fits. Rather, our hope is that people with access to much more information than us (for example the public health authorities of a university or a city) will be able to incorporate their knowledge into our model to obtain case trajectories with high accuracy as well as predictive power.

Before beginning the generalizations, we note three features of the baseline equation (4) which represents the word equation (2). The first is that it represents a **clean decoupling** of the various classes of effects – public health terms come into the first term on the RHS, immune response effects into the second term and intrinsic disease characteristics such as asymptomatic fraction and symptomatic latent period into the third term. This is not the case for the S-E-I-R model. This phenomenon we call separation of parameters (which sounds like a conflation of separation of variables and variation of parameters [24]). This parameter separation means that the question we posed in §2 about the effects of changing the asymptomatic infection period from 4 days to 7 days can be answered in one line basis of (4) [our second question is more involved and the answer is coming later in this Section].

The second noteworthy feature of the baseline equation is that it is cast in terms of a **single variable** rather than an array of variables. Quantities such as the numbers of hospitalizations and deaths can be easily extracted from the cumulative case trajectory by using the known hospitalization and mortality rates and intervals between contraction and recovery or death. Thus, the smaller number of variables does not amount to a limitation on the information that can be obtained from our model. Rather, by using DDE instead of ODE, it is possible to create a versatile model with a less complex appearance.

The third feature of (4) is its **negligible computational cost**. The code for solving (4) takes about 100 lines of Matlab to write, and about one second on a laptop to run. The sizes and runtimes will remain of the same order for all the variations we shall consider in this Article. With this, we go on to our consideration of the individual public health intervention steps.

### §5 AGE/VULNERABILITY AND TRANSMISSIBILITY STRUCTURING

Our first generalization deals with structuring. The most common structuring is age-structuring which is important because the effects of COVID-19 on young and old people are vastly disparate (the latter are much more severely affected on average). We here consider a stratification of society into two classes – young people and old people. To be more accurate, the “young” class includes everyone who is less vulnerable, irrespective of age (for example, immunocompetent 60-year olds with no known comorbidities also qualify) while the “old” class include everyone who is more vulnerable (including say 20-year olds with known immune disorders). Let *y*_1_ denote the cumulative case count in the young population and *y*_2_ the case count in the old population. Instead of a single interaction rate *q*_0_, we now need three interaction rates : *q*_1_, that of young people with other young people, *q*_2_, that of young people with old people and *q*_3_, that of old people with old people. Let *N*_1_ and *N*_2_ denote the initial susceptible numbers of young and old people. Writing (2) for the young people and expressing it mathematically as in (4), we get

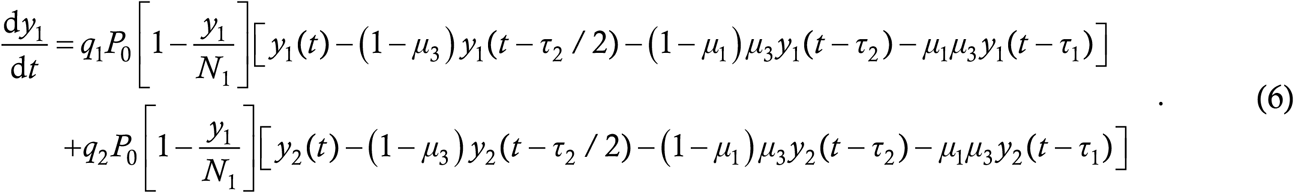

Recall that *P*_0_ is the probability that an interaction between a case and a susceptible target actually results in a transmission. The first line on the above RHS denotes the transmission to young targets from young cases while the second line denotes transmission to young targets from old cases. Note that the susceptibility probability is 1 − *y*_1_/*N*_1_ in both terms – since *q*_1_ and *q*_2_ already accommodate the fact that the target is a young person, the thing to ask here is, given that the target is young, what is the probability that s/he is not immune. By analogy, the case counts among the old people will be given by

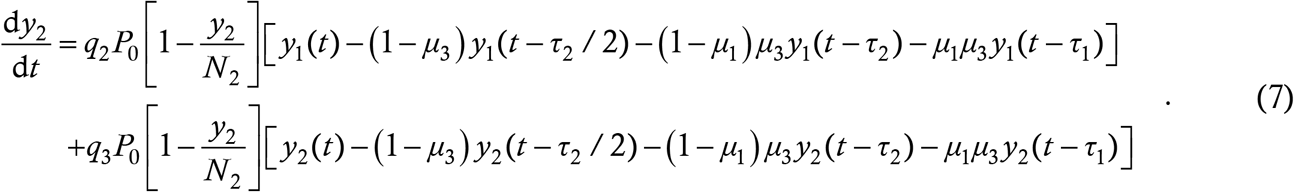

It should be obvious how the model can be generalized to include many more vulnerability classes. Most existing ODE disease models can be extended for age structuring however (the logistic model is a notable exception), so we do not tout this as a signal virtue of the new DDE model.

A structuring which is trickier to handle is the following : suppose that among the *N* interacting people, *N*_1_ wear masks all the time and *N*_2_ don’t wear them at all (*N*_1_+*N*_2_ = *N*). This kind of situation is very common especially in USA where there is considerable resistance to mask-wearing in some places. A structured model is superior to a univariate model here for two reasons : (*a*) it can enable public health authorities to quantitatively determine how much better the mask wearers will fare and accordingly formulate public policy, and (*b*) it is possible that mask wearers might suffer less severe symptoms due to receiving smaller viral loads, so the hospitalization and mortality characteristics might be different among the two groups (whether this is true or not for COVID-19 is currently unknown – a recent study [25] reports a small but statistically significant correlation between mask mandates and hospitalization rates, and we haven’t seen anything more definitive).

The raw input for this situation is a set of four probabilities : the probability *P*_1_ that the disease jumps from a masked case to a masked target when they interact, the probability *P*_2_ that the disease jumps from a masked case to an unmasked target, the probability *P*_3_ that the disease jumps from an unmasked case to a marked target and the probability *P*_4_ that the disease jumps from an unmasked case to an unmasked target. Common sense says that *P*_1_ will be the smallest and *P*_4_ the greatest of the four probabilities; studies seem to indicate that *P*_2_ is less than *P*_3_ i.e. one mask between two people is more useful when on the case than on the target. We assume that the interaction rate is constant i.e. every person interacts at the same rate *q*_0_ with other persons, whether masked or otherwise. We also ignore age-structuring. This last statement is a general philosophy we shall adopt throughout this Article – **when considering each new situation, we shall incorporate the modification directly into the baseline model and not a variant form**. Whoever needs an equation with both age and mask structuring can derive it by him/her-self.

To model this situation, let *y*_1_ denote the cumulative number of cases in the masked population and *y*_2_ the same thing in the unmasked population. Considering the *y*_1_-dynamics first, there will be two terms just as in (6) to account for transfers to masked targets from masked and unmasked cases. For each term, we again have the structure (2). By our assumptions, masked cases interact with all others at an average rate *q*_0_, and since all interactions take place equally, a fraction *N*_1_/*N* of these will be with masked targets. Then, we have the probability *P*_1_ of mask to mask transmission, the probability 1−*y*_1_/*N*_1_ of masked target susceptibility and a set of delay terms for the number of masked cases at large. For the second term, unmasked cases again interact with everyone at rate *q*_0_ and fraction *N*_1_/*N* of these interactions are with masked people. Then we have the probability *P*_3_ of transmission from unmasked case to masked target, followed by the susceptibility probability as above and a second set of delay terms counting the unmasked cases at large. This implies

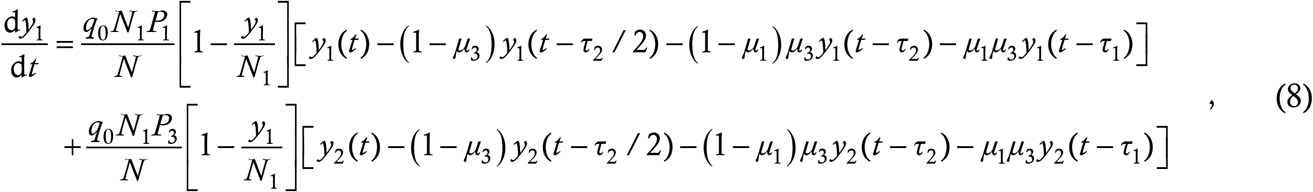

and by analogy

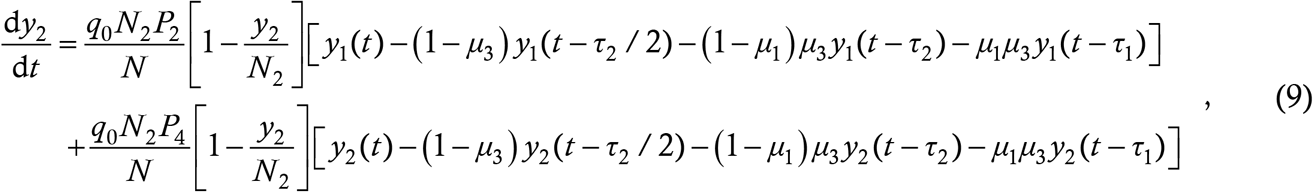

which complete the formulation of the transmissibility-structured model.

Below we show a sample plot of the solutions of (8,9) for the parameters *τ*_1_ = 7 and *τ*_2_ = 3 (as for all simulations in this Article), *μ*_1_ = 0·8, *μ*_3_ = 0·75, *q*_0_ = 4, *P*_1_ = 1/30, *P*_2_ = 1/20, *P*_3_ = 1/15, *P*_4_ = 1/3, *N*_1_ = 2,50,000 and *N*_2_ = 50,000. In these and similar plots, we use the same colours for *y*_1_ as used for *y* in Figure 1, and red for cases, magenta for derivative and cyan bars for epi-curve of *y*_2_.

**Figure 1:**
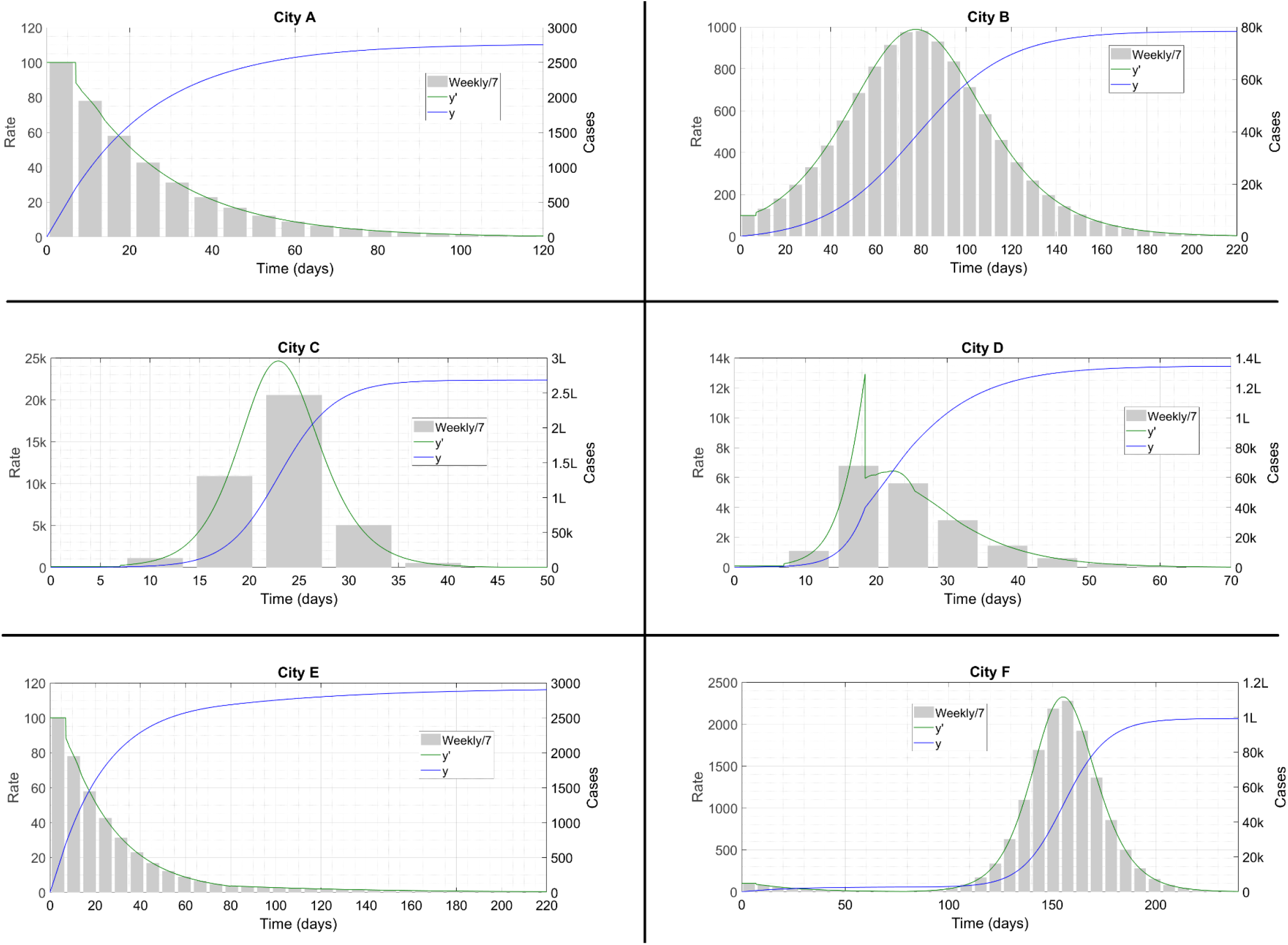
Case curves for the solution classes mentioned in Table 1. Adapted from Reference [20]. The symbol ‘k’ denotes thousand and ‘L’ lakh or hundred thousand.

**Figure 2:**
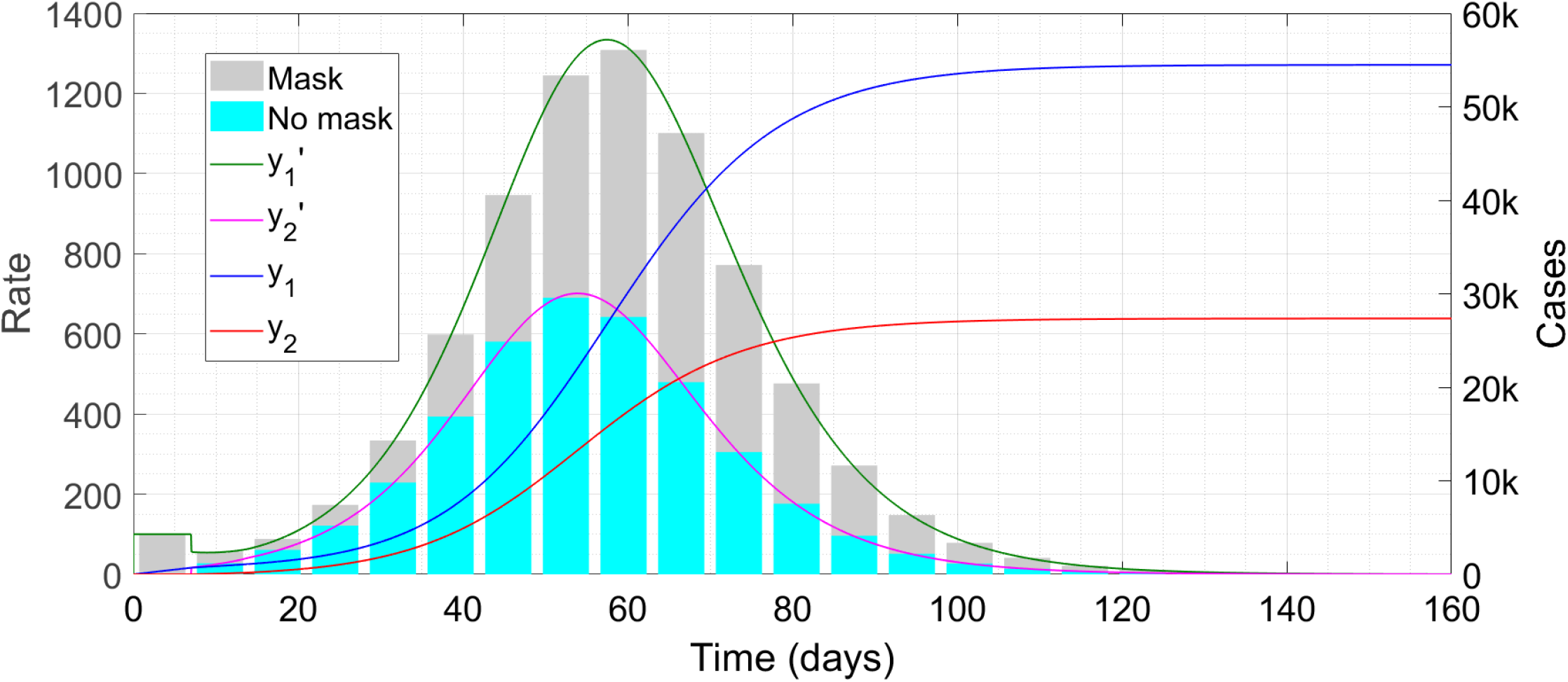
Case trajectories with extensive mask use. The symbol ‘k’ denotes thousand.

**Figure 3:**
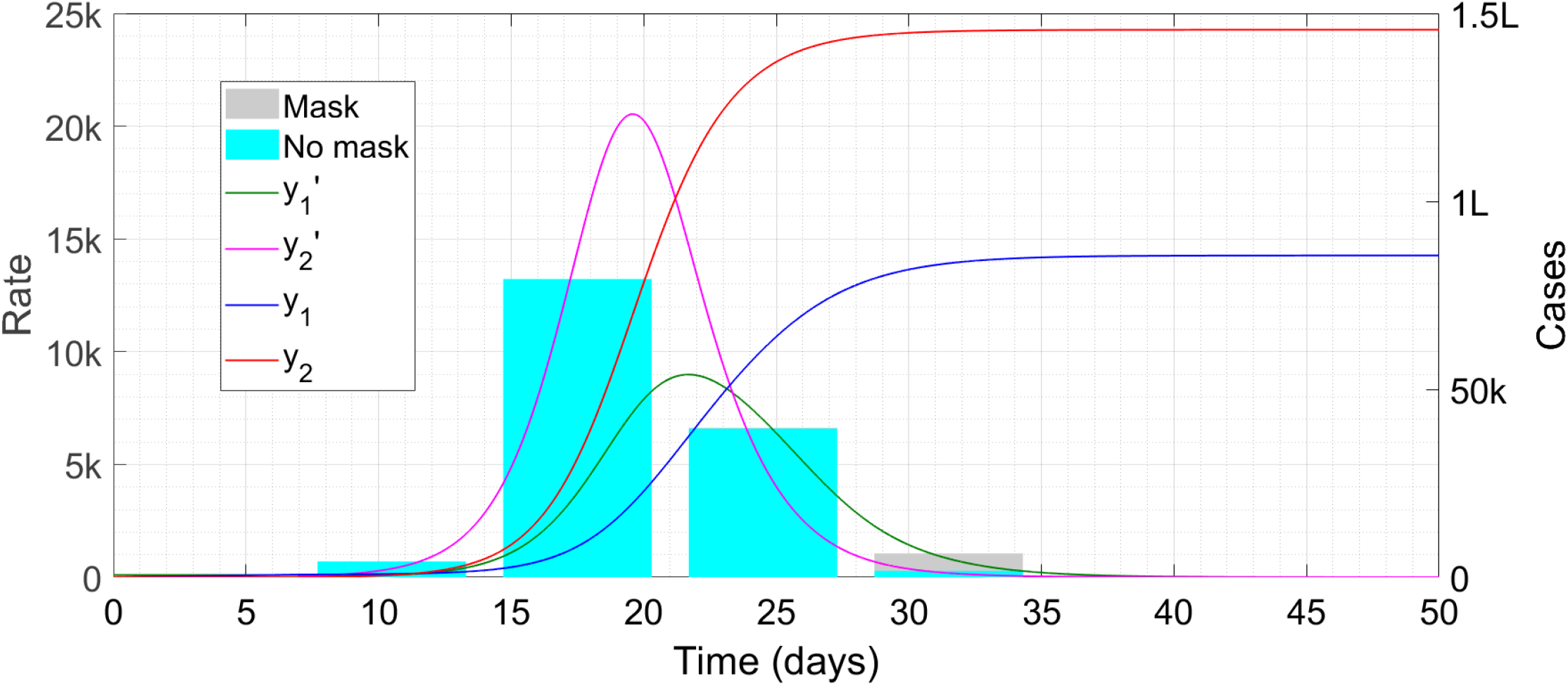
Case trajectories with 50 percent mask use. The symbol ‘k’ denotes thousand and ‘L’ lakh or hundred thousand. Note that the cyan bars obscure the grey bars in many places; the envelope of the latter is the green curve which is still visible.

We can see about 55,000 cases among the masked population which amount to just above 20 percent of the total 2,50,000 masked people. By contrast there are about 27,000 unmasked cases which amount to more than 50 percent of the 50,000 unmasked people.

**Table 1:** Six solution classes of the baseline model (4). These correspond to different kinds of corona trajectories seen all over the world.

| City | $m_0$ | $\mu_3$ | Description | Example |
| --- | --- | --- | --- | --- |
| **A** | 0.23 | 0.5 | Epidemic contained via public health measures | All cities in New Zealand |
| **B** | -do- | 0.75 | Epidemic grows initially, reaching partial herd immunity [20] | Most or many cities in India and in California (USA) |
| **C** | 0.5 | -do- | Epidemic progresses to herd immunity with little intervention attempted | None |
| **D** | 0.5 upto 40,000 cases and 0.23 thereafter | -do- | Late lockdown causes initial explosion before settling down to partial herd immunity | New York City, USA |
| **E** | 0.23 upto 80 days and 0.5 thereafter | 0.5 upto 80 days and 0.1 thereafter | Advanced contact tracing drive enables reopening with no spreading | None |
| **F** | -do- | 0.5 upto 80 days and 0.2 thereafter | Reopening causes a second wave | Cities in UK, Germany |

We now run the code again, this time setting *N*_1_ = *N*_2_ = 1,50,000 i.e. equal numbers of masked and unmasked people.

Not only is the unmasked population infected almost entirely but also there is more than 50 percent infection level among the masked people. This type of statistic can be used by public health authorities to encourage mask use – by not masking, not only are you increasing your own chances of catching corona but you are subjecting the law-abiding people to extra risk as well. Of course, for a propaganda drive, we first need good-quality data on the efficacy of mask use – the values of *P*_1_ to *P*_4_ we used here for the simulations were just rabbits taken out of a hat. Two sources which can yield these values are (*a*) laboratory experiments and simulation studies, and (*b*) follow-ups of exposures arising from contract tracing activities, including cases as well as non-cases who were exposed to a known transmitting case.

Transmissibility structuring can also be used to model the effect of super-spreaders, who transfer the disease to many targets. Super-spreaders can be of two kinds – people who interact with others a lot, and people who have exceptionally high viral loads and infect almost whomever they come into contact with. To model this setup, we can define two classes of people, superspreaders and “normal” spreaders, with the former having smaller population and higher interaction rates and/or transmission probabilities compared to the latter. Since the equations for this situation will show almost no difference from the mask structured model, we dispense with a detailed derivation.

### §6 CONTACT TRACING

This is one of the more difficult aspects to incorporate into a lumped parameter model, and is an area where agent-based models have an intrinsic advantage. The DDE however is capable of taking this in its stride. As we have already seen, the baseline model itself incorporates contact tracing to a fair extent. It contains two assumptions however – those of zero non-transmissible incubation period (NTI) and instantaneous tracing – which cannot be included in a more detailed model where contact tracing is a priority. It also contains a heuristic parameter *μ*_3_ which is too coarse for the present application.

To develop this aspect, we first assume that there is no contact tracing whatever; then (4) looks like

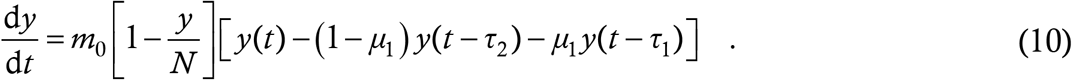

Now we have to throw away the assumption of zero NTI. For this, we introduce a new variable denoting the **exposed** people. In the ODE models it is denoted by *E*; since we are using *y* for cases, we see no harm in using *x* for exposures instead (in general we prefer *x* (*t*) to *y* (*t*) for a single variable – the nomenclature here is a carryover from our original somewhat cumbersome four-variable model [26] where *x* denoted susceptible people, *y* infections and so on). We define that a person transitions from *x* to *y* at the instant s/he turns transmissible, which is also the moment when s/he will first return positive if tested for the virus.

What the RHS of (10) denotes is actually the rate of exposures – interaction between cases and targets leads to exposures rather than cases among the latter. So, we can replace the left hand side (LHS) of (10) by d*x*/d*t* as a first step in going past the zero NTI assumption. Doing this we have

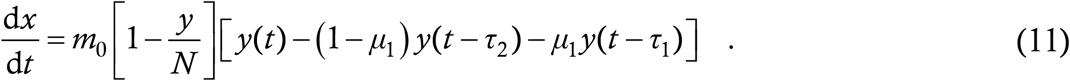

Now let *τ*_4_ denote the period during which a person remains exposed before turning into a case – as in all lumped parameter models, this must be an average over all cases. Then, all the people who got exposed

between time *t* and *t* + Δ*t* will turn into cases between *t* + *τ*_4_ and *t* + *τ*_4_ + Δ*t*. This implies that 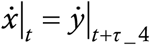 or,

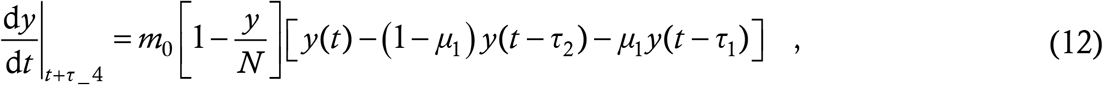

or,

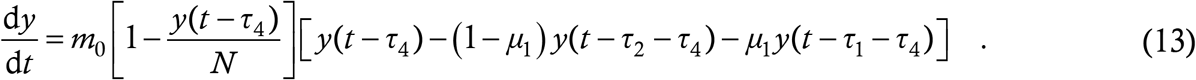

The difference between (10) and (13) is just a shift of the infection curves by *τ*_4_, which is a small quantity in relation to the epidemic’s overall progress (a typical value of *τ*_4_ is 4 days). Hence, in the baseline model we could afford to make the zero NTI assumption without significant error.

For a sophisticated contact tracing model however, a little reasoning shows that this assumption is unacceptably simplistic. For if a patient’s contacts were to be rounded up as soon as they have been exposed, then they would all go into quarantine during their NTI itself and not spread the disease to anyone. In practice however, contact tracing is time-consuming – the starting patient’s test results have to come in (which itself may take a day or more), authorities have to talk to the patient, find out the names and places he gives, call the people involved, access CCTV footage and/or card histories in public places (if permitted) etc. The authorities might be understaffed/overworked and might take time to get to the patients; the patients might not remember their entire movements accurately and might give out the story piecemeal. So, by the time the authorities get to the contacts, some will have already crossed the NTI into the transmissible asymptomatic or latent infection stage.

We assume here that the contact tracing takes place as follows. When a symptomatic patient reports corona to the authorities, they start identifying the people with whom s/he came into contact over the past *τ*_2_ days. They take a time *τ*_3_ to unearth and quarantine a fraction *P*_3_ of the cases among these contacts. The quarantine of healthy people is an unpleasant collateral of a contract trace and does not affect our model. With this strategy of one-level, forward contact tracing, let us try to formulate the equivalent of (11). The heuristic contact-traced fraction *μ*_3_ of (4) must now get replaced by an expression cast in terms of the ground realities *τ*_3_, *τ*_4_ and *P*_3_ and the other existing parameters.

When a symptomatic case, say Mr X, just reports for quarantine, he will have spent the preceding *τ*_2_ days unknowingly spreading virus to targets. In any lumped parameter formulation, we must assume that Mr X spreads the disease uniformly and continuously during his latency period – this assumption makes sense when not one but a hundred simultaneous cases are taken into consideration. Among Mr X’s targets are the person Ms Y he infected right when he turned transmissible and the person Ms Z he infected just before reporting for quarantine. When Mr X locks himself up, the former has spent *τ*_2_ days at large while the latter has spent 0 days at large. The authorities take a further *τ*_3_ days to obtain these contacts, at which point Ms Y has got an exposure time of *τ*_2_+*τ*_3_ while Ms Z has got an exposure time of *τ*_3_. There are three possible scenarios now.

#### Case 1

*τ*_4_ > *τ*_2_+*τ*_3_. In this case, Ms Y is still in NTI and hence every other contact of Mr X is in NTI as well. Those that are identified do not transmit – the average time *T* spent transmissible and at large by the contact traced cases is identically zero.

#### Case 2

*τ*_2_+*τ*_3_ > *τ*_4_ > *τ*_3_. In this case, Ms Y has passed NTI period into transmission but Ms Z is still in NTI. Let *t* = 0 be the time when Mr X turned transmissible so that *t* = *τ*_2_ is when he quarantines, and let *n* be the total number of contacts whom he has infected with *r* = *n*/*τ*_2_ being the infection rate. By hypothesis, the contact trace occurs at *t* = *τ*_2_ + *τ*_3_; the last of the exposures who turns transmissible before being traced is the one who was exposed at *t* = *τ*_2_ + *τ*_3_ − *τ*_4_. The target who was exposed to Mr X at any time *t* prior to *τ*_2_ + *τ*_3_ − *τ*_4_ turns transmissible at time *t* + *τ*_4_ and thus remains transmissible and at large for a duration (*τ*_2_ + *τ*_3_) − (*t* + *τ*_4_). The total time which all these cases have spent at large is

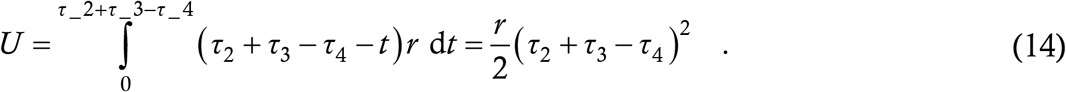

Recalling that *rτ*_2_ = *n* and dividing by *n* gives the average time spent at large by contact traced cases as

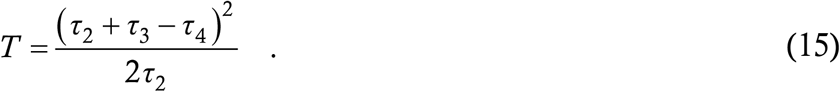

If *τ*_3_ and *τ*_4_ are both zero (which marginally satisfies the conditions of this case), i.e. we make the assumptions of zero NTI and instantaneous tracing, then this reduces to *τ*_2_/2 as it had better.

#### Case 3

*τ*_4_ < *τ*_3_. In this case Ms Z turns transmissible as well before isolation. We can use a procedure similar to Case 2 above to calculate an average time for which the contact traced case remains at large. Since the situation is implausible and the algebra is tedious, we skip this exercise. If teaching this model in a class, we will assign this as a hacking-type homework assignment or exam question.

So we have found the average duration *T* that a contact traced case remains at large – what we now need is the absolute probability that a random case is contact traced (recall that *P*_3_ is the **conditional** probability that a case is contact traced, **given that** s/he was exposed by a symptomatic case). This absolute probability is composed of two probabilities : (*a*) that the random case has been infected by a symptomatic case and not an asymptomatic one (secondary cases of asymptomatic cases do not get picked up by definition), and (*b*) *P*_3_. To calculate the probability (*a*), we use a self-consistency procedure.

Let this unknown probability be *X*. We assume, very plausibly, that the secondary contacts of every case (symptomatic or otherwise) are asymptomatic with the fraction *μ*_1_ (as defined for the disease as a whole) and symptomatic with the fraction 1−*μ*_1_. Consider a random asymptomatic case. With probability *X* she has been infected by a symptomatic case; with further probability *P*_3_ she gets picked up in the contact tracing drive. If she does, i.e. with probability *P*_3_*X*, she spends time *T* at large. Otherwise, she remains at large for the entire asymptomatic infection period *τ*_1_. Combining the two, on average an asymptomatic case remains at large for a time

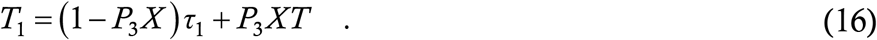

Similarly, a symptomatic case gets picked up in the drive and spreads for time *T* with probability *P*_3_*X*; he spends the entire latency period *τ*_2_ at large otherwise. Thus, the average time spent at large by symptomatic cases is

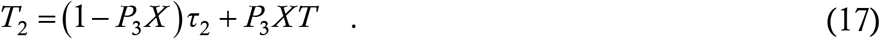

So we have a fraction *μ*_1_ of asymptomatic cases remaining at large for time *T*_1_ and a fraction 1−*μ*_1_ of symptomatic cases remaining at large for time *T*_2_. Since by the model assumptions all cases transmit equally and uniformly, the total numbers of secondary cases spawned by these two types of cases must be in the ratio *μ*_1_*T*_1_ to (1−*μ*_1_)*T*_2_. This ratio must be **the same as the ratio of the probabilities** that a random case has been infected by an asymptomatic and a symptomatic case respectively; thus we have

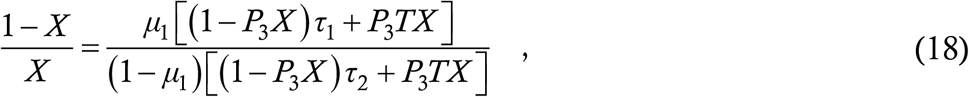

which rearranges into a quadratic equation for *X*

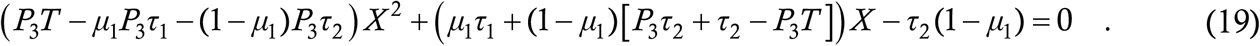

One of its two roots, *X* = *X*\*, should be a number between 0 and 1, which is the desired probability.

Finally, we are at a stage where we can write the model equation. A random case is contact traced with probability *P*_3_*X*\*, so this is the equivalent of 1−*μ*_3_ in the baseline model (4). The contact traced cases remain at large for duration *T* as calculated in the case-wise analysis. Thus, the exposure equation is given by

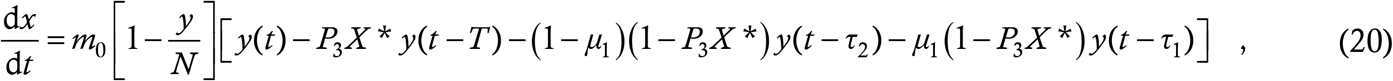

and the case equation follows as

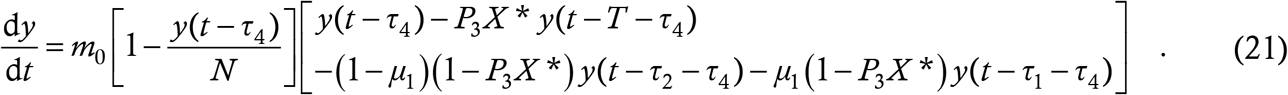

The structure is identical to (13) and very similar to (4) but the terms *T* and *X*\* now arise from calculations of considerable sophistication, based directly on the details of the contact tracing process.

Similarly, it should also be possible (though hasslesome) to implement two-level contact tracing where contacts of contacts are also identified and grounded. We leave this task to public health authorities who are actually carrying out contact tracing operations (or to students of a disease modelling course who are willing to take on pathologically long assignments just for a few more marks).

Since (21) has nearly the same form as the baseline, we refrain from showing simulation time traces. Rather, we give the values of *X* and hence of the capture probability *P*_3_*X* or 1−*μ*_3_ for some realistic sets of parameter values. With the standard values 7 and 3 for *τ*_1_ and *τ*_2_ and taking *τ*_4_ = 4 (the total incubation period is approximately 7 days on average [21]), the value *τ*_3_ = 2 gives *T* = 1/6. This implies that contact traced cases remain at large for a relatively short time. The fraction of cases which is caught depends highly on two parameters – the asymptomatic fraction *μ*_1_ and the tracing success probability *P*_3_. Thus, if 30 percent cases are asymptomatic, then *P*_3_ = 1/2 leads to capture of 25 percent of the total cases while *P*_3_ = 9/10 leads to capture of 46 percent of the total cases. If 80 percent cases are asymptomatic however, then *P*_3_ = 1/2 captures not even 5 percent of the total cases, which increases to 8·7 percent if *P*_3_ is raised to 9/10.

This is not a surprise. Since the contact tracing is being carried out starting from symptomatic cases, the process efficiency automatically becomes crippled if they are few in number. Tracing tertiary contacts i.e. contacts of secondary cases will not really help since the secondary cases get quite a low air-time anyway. Two-way contact tracing, i.e. when a new case reports symptomatic then trying to identify the case’s source of exposure, and again following up on that source’s other exposures can lead to the capture of many more cases. This process is difficult however since an exposure typically occurs 7 days prior to symptoms, and a typical patient will hardly be expected to remember his/her movements from 7 days past. Another way of capturing asymptomatic cases is by performing regular mass testing; this we discuss below.

### §7 MASS TESTING

Periodic testing of the entire population is yet another disease mitigation strategy which is employed in organizations with small staffing and large funding. The White House, USA is one example. Another example is the Ithaca campus of Cornell University, USA where one of us (SHAYAK) is located. Cornell tests almost the entire campus community once a week for the virus (and also employs a battery of surveillance and punitive measures to prevent student misbehaviour). After an initial outbreak (followed by harsh crackdowns), the number of on-campus cases here has remained very low, with every week seeing single-digit new case counts.

To model this effect, we start as usual from the underlying structure (2). As with contact tracing in the last Section, the first two terms remain unchanged – what frequent testing changes is the duration for which cases remain at large. To lend concreteness to the discussion, we introduce numerical values at this step itself. Let the testing be carried out once a week, let the test itself be perfectly sensitive and let the results take one day to come through. The asymptomatic and latent periods remain *τ*_1_ = 7 and *τ*_2_ = 3 as they always were. Let Mr X be an asymptomatic case who does not get contact traced and has his test on a Monday. With equal probability 1/7, Mr X can turn transmissible on any day of the week. If he starts on a Monday, he transmits for only one day before he is detected and quarantined. If he starts on a Sunday, he goes 2 days at large before capture, if Saturday then 3 days and so on until if he turns transmissible on a Tuesday, bad luck, he spends the entire 7-day asymptomatic infection period at large. The average duration for which he remains at large is (1/7) (1+2+3+4+5+6+7) which is 4 days. Similarly let Ms Y be an untraced symptomatic case who gets tested on Monday. She transmits for 1 day if she starts on a Monday, 2 days if Sunday and the full latency period of 3 days otherwise. Thus, the average duration for which she remains at large is (1/7) (1+2+3×5) which is 18/7 days.

For contact tracing, we go back to the simple structure of the baseline model instead of using the complications of the last Section – a heuristic parameter *μ*_3_ for the escape fraction and the assumptions of zero NTI and instantaneous tracing. This time however, tracing will proceed from **every** case which is picked up during testing. The traced cases will spend an average at large time of half their sources; these latter spend 4*μ*_1_ + (18/7) (1−*μ*_1_) days. Thus we have the model

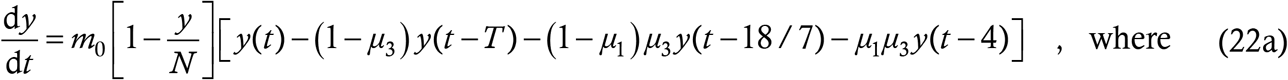

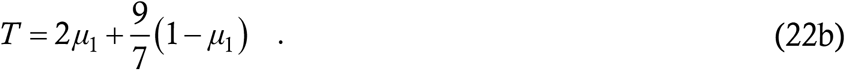

Once again, we have the same basic structure as (4) with new parameter values. This answers the second question with which we had challenged the S-E-I-R model in §2.

It is also possible to incorporate the effect of imperfect test sensitivity without too much fuss. A study [27] indicates that the “gold standard” RT-PCR test has a sensitivity of approximately 75 percent or lower; the antigen tests are typically less sensitive. With imperfect sensitivity, if a person is a case, the test has a probability *P* of actually reporting positive and a probability 1−*P* of coming out false negative. We assume that the tests are fully specific i.e. there are no false positive cases (this assumption is consistent with reality). To calculate the durations for which cases remain at large, we use the same argument as above. For the asymptomatic Mr X, this time there is a probability *P* that he gets caught after 4 days and a probability 1−*P* that he remains at large for the full 7 days. Similarly for the symptomatic Ms Y, there is probability *P* of capture after 18/7 days and probability 1−*P* of remaining at large for 3 days. The asymptomatic at large time is therefore

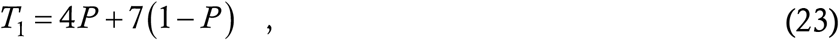

while the symptomatic at large time is

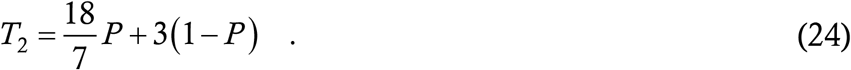

Using these in the basic structure (2) we have

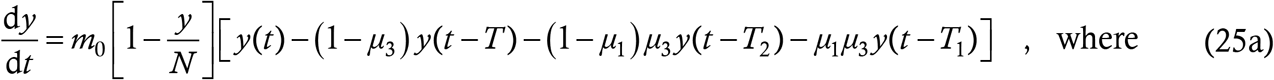

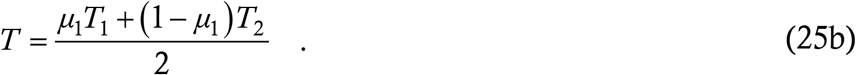

Thus, another sophisticated effect, namely mass testing with limited sensitivity, has also become incorporated into our DDE model.

Since the equation (25a) is similar in structure to (4), we again refrain from showing simulations. This time, a relevant question is, what is the effect of the testing drive on the reproduction number *R*. Repeating the procedure [22] which led to (5) shows that *R* for (25a) is given by

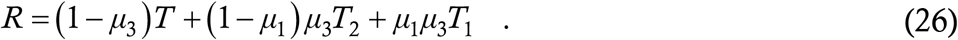

Using the standard values of *τ*_1_ and *τ*_2_, *μ*_1_ = 0·8 and *μ*_3_ = 0·9, we calculate the reduction in *R* if the weekly testing drive is implemented while keeping everything else the same. We find a 38 percent reduction if the test is 100 percent sensitive and a 22 percent reduction if the test is 60 percent sensitive. If *μ*_1_ = 0·3 and *μ*_3_ = 0·5, then the reduction drops to 21 percent with fully sensitive test and 8·5 percent with 60 percent sensitive test. The decrease from the previous situation is not surprising since the weekly testing does not significantly reduce the at large duration of symptomatic cases. The value of *μ*_1_ = 0·3 is too low however and the earlier set of figures is more realistic. A 20-30 percent reduction in *R* is not jaw-dropping but not trivial either – it is ineffective against a really high *R* of say 2 but it can bring down a somewhat-above-unity *R* to just below unity and extinguish the epidemic in time. This is more or less what is happening at Cornell University – the constant threats of disciplinary action are ensuring that students are not driving up the default *R* to an unmanageable value, and thereafter the testing program is keeping it further down.

Again we note that although the interventions of masking, contact tracing and testing are all currently relevant in the context of corona, they are applicable to other infectious diseases as well as such as pandemic influenza and will very likely be used whenever a new infectious epidemic breaks out anywhere in the world.

## 3 IMMUNE RESPONSE EFFECTS

While the last Part dealt with public health interventions, in this Part we turn to another aspect of the disease which is the human immune response. The baseline assumption is that of permanent immunity – one bout of infection renders a person insusceptible to the disease for all time. This assumption is valid whenever the immunity period is longer than the overall progression of the epidemic – immunity need not be genuinely lifelong. For corona, about half a dozen phylogenetically confirmed cases of second time infection have been found worldwide so far – 1 in Hong Kong, 2 in India, 2 in the US, 1 in Belgium, 1 in Ecuador and there may be a couple more which we may have missed. Less convincing evidence of reinfection exists for a few more cases, although the overall fraction of reinfections is negligible as least yet (and we hope it stays that way). A large scale antibody study [28] has found that detectable antibody titres persist in blood for at least three months following infection; we are yet to see a follow-up to that study. The question naturally arises as to what profile the spreading trajectories may take if immunity really turns out to be temporary in a significant fraction of the population. Once again, our focus here will not be on an exhaustive study of the results in various cases but on how we can incorporate different kinds of immune response into our DDE model.

### §8 SIMPLE TEMPORARY IMMUNITY

Here we consider the case where immunity against the disease lasts for a fixed, limited duration following the first infection. After this duration is over, the person becomes susceptible to the disease again. One study [29] finds such a situation for benign human coronaviruses (**not** SARS, MERS or COVID-19). Let *τ*_0_ be the duration for which immunity lasts. As is customary in lumped parameter models, the value we use must be the average over the entire population.

As usual, we start from the word equation (2). Here, the second term on the RHS denotes the immune response so only that will be altered relative to the baseline model (4). The term must still denote the probability that a random person is susceptible. By our assumptions, each recovered case remains immune or insusceptible for a duration *τ*_0_, the immunity cutoff period, typically of the order of weeks or months. So, if a person once contracts and recovers from the infection at time *t*, then s/he remains immune upto time *t* + *τ*_0_ and then becomes susceptible again. At any given instant, the people who are immune are all those who have contracted the infection during the last *τ*_0_ days, and no one else. Hence, the number of insusceptibles at time *t* is exactly the number of new infections which have occurred between time *t* − *τ*_0_ and *t*, which is *y* (*t*) − *y* (*t* − *τ*_0_). The structure of this term and the underlying logic are the same as those motivating the other delay terms in (4). In proposing this structure, we have again assumed that recoveries are instantaneous (see §3) and also have ignored deaths. Since the mortality rate of COVID-19 is fortunately quite low, this second assumption is reasonable as well. Then, the probability that a random person at time *t* is susceptible is

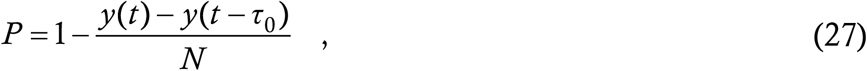

and working this into (2) yields

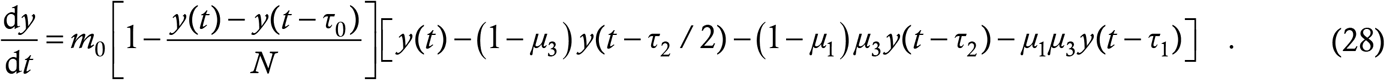

This is the generalization of the baseline model to the case where immunity is temporary.

We run a simulation with parameters *m*_0_ = 0·23, *μ*_1_ = 0·8, *μ*_3_ = 0·75 and *τ*_0_ = 200 days. The result is shown in Figure 4.

**Figure 4:**
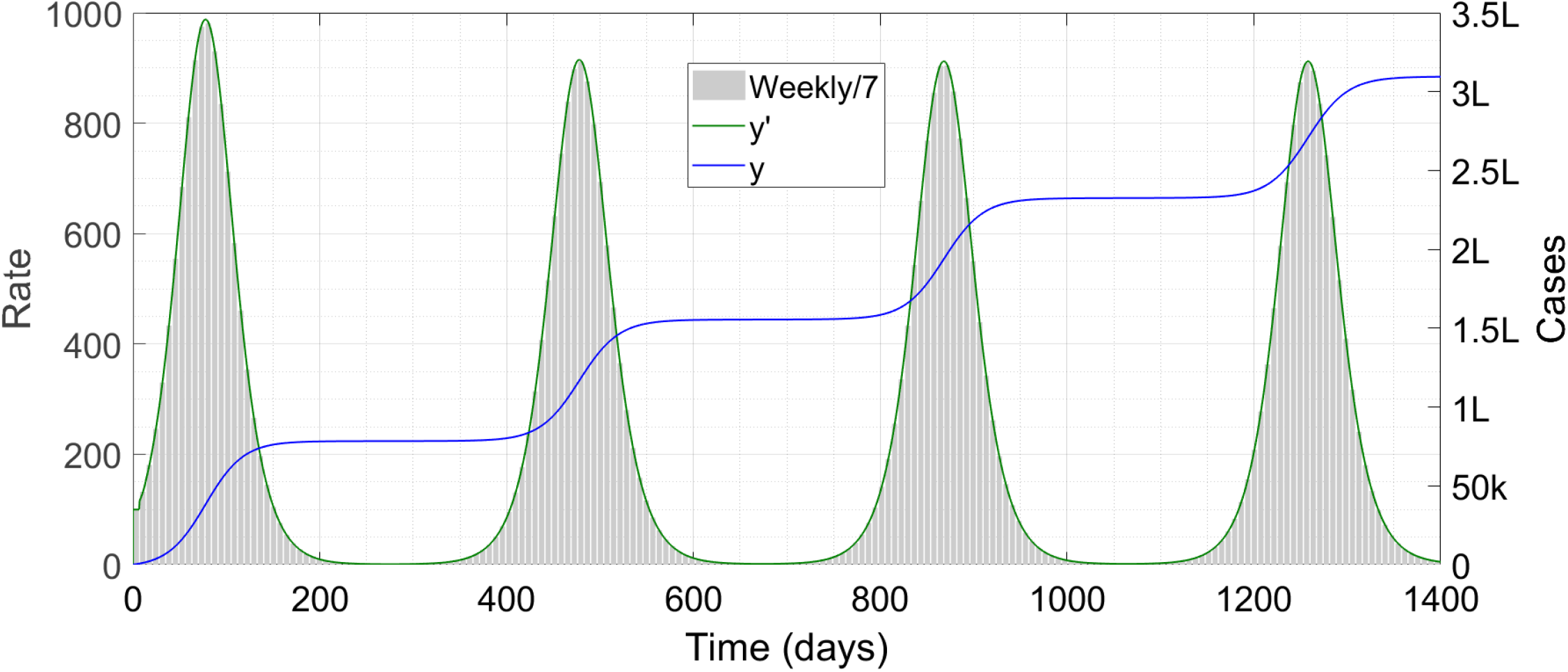
Case trajectories with temporary immunity. The symbol ‘k’ denotes thousand and ‘L’ lakh or hundred thousand.

We can see multiple waves of disease in this case, which is consistent with what has been found in the prior literature [30,31] using different models. Note that the waves continue indefinitely; we have artificially stopped the simulation at *t* = 1400 days.

### §9 COMPLEX IMMUNE RESPONSE

The assumption that every case becomes completely susceptible after a certain time is perhaps over-simplified. Specially for COVID-19, we do not have nearly enough data about the immune response to the pathogen. However, for diseases in general, there are three kinds of immunity – sterilizing immunity which completely prevents reinfection, severity-reducing immunity which mitigates the symptoms during reinfection and transmissibility-reducing immunity which mitigates the patient’s transmissibility during reinfection. Studies on the benign coronaviruses [32] have found that although sterilizing immunity wanes in about a year, severity-reducing immunity lasts much longer. In addition to the above, there is a dangerous phenomenon called antibody-dependent enhancement (ADE) in which case a reinfection takes a more severe form than the original infection. This happens as a result of imperfect binding of antibody to the virus, followed by a misrecognition of live virus for inactivated virus by the immune system. For certain viruses like dengue virus, ADE can happen as an intrinsic phenomenon in almost all patients; for other pathogens, some patients can still experience ADE due to immune system malfunction. ADE has not been found for benign coronaviruses; it is unknown whether it occurs with COVID-19. Among the six genetically confirmed reinfection cases so far, two reported a more severe infection the second time around while the others reported milder infection. It is thus possible that a mixture of the various immune responses are occurring with COVID-19 – time alone will throw further light on this matter.

We consider a situation where every recovered case has the probability *P*_1_ of remaining immune for life, *P*_2_ of remaining fully immune for *τ*_0_ days and then becoming susceptible to a lower virulence form (LVF) of the disease and *P*_3_ (with *P*_1_+*P*_2_+*P*_3_ = 1) of remaining fully immune for *τ*_0_ days and then becoming susceptible to a higher virulence form (HVF) of the disease through ADE. We further assume that after catching the infection twice, a person does not contract it any longer (becomes either immunized or dead). To model this situation, we define three variables *y, z*_1_ and *z*_2_. Here *y* denotes the count of cases in the current form of the disease, *z*_1_ denotes the count of LVF cases and *z*_2_ the count of HVF cases.

We start from the *y*-equation. Any person who catches the current form of the disease once is insusceptible to it for all future time – s/he contracts either nothing, or LVF, or HVF. Thus the susceptibility probability in this equation will be 1 − *y*/*N* as in the baseline model. As in the coupled models of §5, the number of at large cases will include all the forms of the disease, so there will be three sets of delay terms. We assume, for the sake of notational elegance more than anything else, that LVF and HVF have the same infection parameters as the current form. We then have

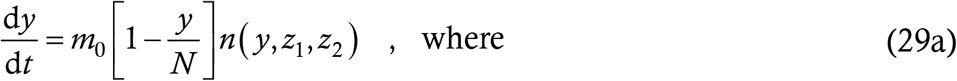

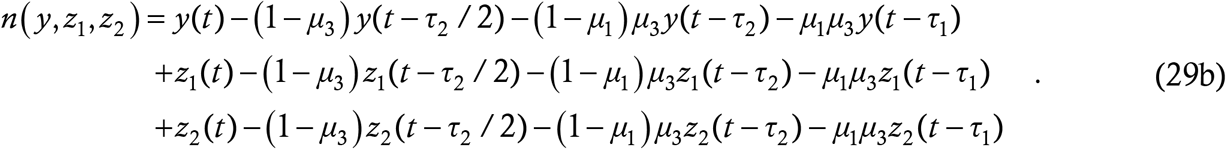

The notation *n* (*y,z*_1_,*z*_2_) is just a shorthand which shall save us the trouble of typing a repetitive three-line expression two more times.

For the *z*_1_-equation, we ask the usual question that what is the probability of a random person’s catching the LVF disease. For this we have to first count the number of people who are eligible to catch the LVF. This includes everyone who has already contracted the default form, and that a time *τ*_0_ or more days ago, which is *y* (*t*−*τ*_0_). From this eligible pool however, we shall have to exclude all those who have already had the second infection, either LVF or HVF. So the pool of eligible targets has the size *y* (*t*−*τ*_0_) − *z*_1_(*t*) − *z*_2_(*t*). Given that a person is eligible, the probability of his/her catching LVF is *P*_2_. Putting all this together, the probability of a random person’s being susceptible to LVF is

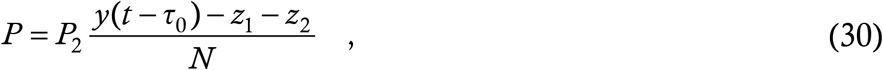

which leads to the equation for *z*_1_ as

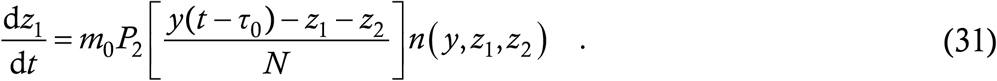

It is now a simple matter to construct the equation for *z*_2_, where everything remains the same as above except that the probability *P*_2_ is replaced by *P*_3_, giving

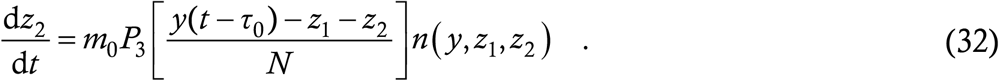

Equations (29,31,32) constitute a coupled set of DDEs for the dynamics of the current and the modified virulence forms.

For the simulation run, we consider the same parameters as in §8 above and take *P*_1_ = 2/5, *P*_2_ = 3/5 and *P*_3_ = 0. Thus, every person who has had one bout is either permanently insusceptible (40 percent chance) or susceptible to the LVF after 200 days (60 percent chance). The run is below, in Figure 5.

**Figure 5:**
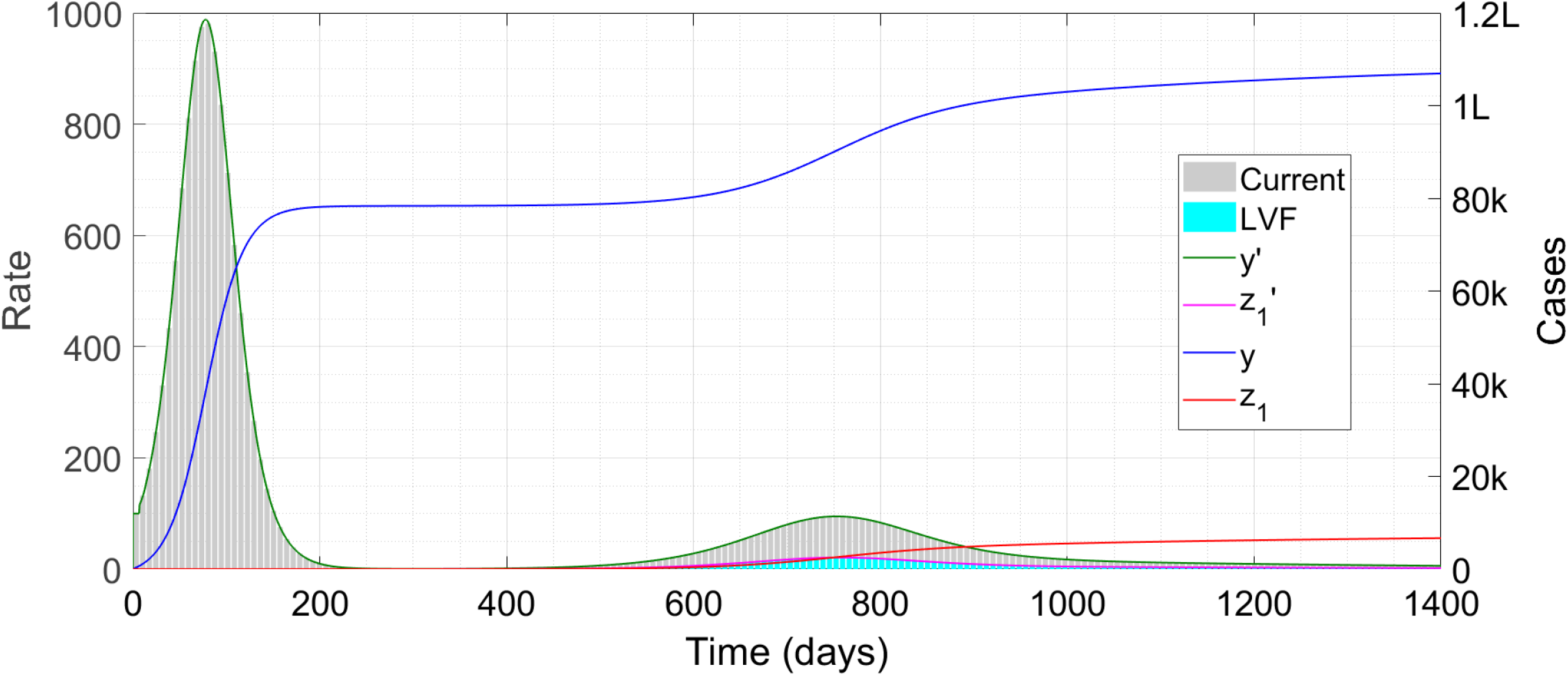
Case trajectories with given probabilities of different kinds of immune response. The symbol ‘k’ denotes thousand and ‘L’ lakh or hundred thousand.

We can see that instead of multiple identical waves as in Figure 4, there is only a very broad and low second wave. The first wave is entirely in the current form of the disease while the second features both the current and the LVF forms. Because the probability of catching LVF is quite low, the wave in that form is even smaller than in the current form. As in Figure 4, the runs continue beyond 1400 days but we have stopped it at that time. The total numbers of *y* and *z*_1_ are bounded by *N* and 3*N*/5 i.e. 3,00,000 and 1,80,000 respectively, which automatically prevents infinite perpetuation of the waves; the wave shapes are modified to fit this constraint.

Most of the assumptions we have made in this Section can be relaxed very easily. Incorporating different transmissibilities and durations for the three forms of infection is trivial, while working round the maximum two infections constraint is also easy. For the latter situation, we introduce more forms of the disease, say ultra-LVF *w*_1_ and ultra-HVF *w*_2_ and ploddingly calculate the susceptibility probability of each person to the various infections to use in the structure (2). We shall not embark on this exercise unless emerging corona data shows it to be necessary.

## CONCLUSION AND FUTURE DIRECTIONS

First we give an outline of this rather lengthy Article. In one sentence we can say that the Article is in passacaglia form. After the Introduction (§1,2), in §3 we have presented a “word-equation” (2) which forms the super-structure for our various models. The unadorned bassline is (4), whose solutions we have presented in §4. These solutions show strong similarities with the actual COVID-19 trajectory data observed in various parts of the world. The next five Sections are the passacaglia itself as we treat the bass to one after another variation. The first variation (§5) is the incorporation of age and transmissibility structuring. The second variation (§6) is a detailed model of contact tracing, a process which is traditionally considered difficult to incorporate into a deterministic lumped-parameter model. The third variation (§7) is weekly testing, which is currently relevant in certain limited contexts but might become more widespread as more and more testing kits are manufactured and/or new testing technologies developed. The fourth variation (§8) deals with simple temporary immunity where a recovered case again becomes susceptible to the disease after a time interval. The fifth and last variation (§9) deals with a complex immune response where a recovered case can show different kinds of responses with different probabilities.

Our study is extremely relevant at the current time. Although corona has been with us a good few months, it is unfortunately far from over. With many countries in the grip of sweeping second waves, a versatile model with all a priori parameters is essential to make predictions of hospitalizations and deaths, both with status quo and with different interventions hypothetically applied or removed. Part 2 covers these situations in detail. Part 3 is a glimpse into a potential future and will become pressingly useful if COVID-19 immunity really turns out to be temporary in a significant fraction of the population. It is eminently possible to create models which combine different variations, for example a model with mask-structuring, realistic contact tracing and complex immune response. Bolstered with good data, such models should have high predictive power. There have been several cases of model failure and consequent public health mistakes with COVID-19; New York State, USA [33] is one of the more conspicuous examples. We hope that our new model, with a simple and robust baseline option and a plethora of customizable augmentation options, shall be more resilient to failures of this kind.

Lumped parameter models do have certain intrinsic limitations. While ours is particularly versatile, there are some places where it nevertheless runs up against a wall. For example, the parameters *q*_0_, *P*_0_ in the baseline model have to be averaged over all at large cases. During contact tracing we were forced to make assumptions regarding continuous and uniform transmission etc. For these reasons, DDE cannot replace agent-based models. There are certain questions which a lumped parameter model can never answer. “Today there are exactly 5 active cases in a locality – what is the probability that three weeks later it will be (*a*) zero ? (*b*) more than 100 ?” is one such question. As noted in §1 however, agent-based models are very demanding in terms of computational time and effort; many such studies [4,34] use networks of 10,000 people and not larger. It is noteworthy that the two works we just cited both report on phenomena which are traditionally considered outside the ambit of lumped-parameter models; containment of the epidemic in the former study and mass testing and isolation in the latter. Our model is able to deliver almost all (an exception being for example the stable linear solution of Reference [4]) the results of the agent-based models at a minuscule fraction of the computational cost.

The versatility and computational ease also mean that any newly reported information regarding say the immune response can be incorporated immediately, and the resulting case trajectories computed virtually on the same day that the study is released. This will be a huge advantage when a vaccine is finally released for mass use. By that time, we shall know much more about the immune response to the disease and to the vaccine than we do now. So, instead of performing advance analyses of vaccination [31] with insufficient information, we can wait till concrete info arrives and then quickly build that into our model. After that we can use it for important calculations such as how to apportion the initially limited doses of vaccine to ensure fastest deceleration of the pandemic, and how many doses need to be administered in what kind of timeframe to eradicate the disease altogether.

Finally, the bulk of this Article deals with the specific disease COVID-19, whether implicitly or explicitly. This is only natural since we are writing at the height of this raging pandemic. Nowhere however has this limited our scope. COVID-19 is a particularly vicious virus so it has probably all the weaponry required to generate a successful pandemic – asymptomatic and latent transmission are two of the most effective ammunitions. Indeed, it is not often that a pandemic can grind the entire world to a halt – the last time this had happened was a century ago with a particularly virulent strain of influenza (good boys and girls don’t name viruses after places). With the advent of modern medicine, a pandemic like this was not something that anyone even dreamt of. Less successful pandemics such as SARS-1 and 2005 bird flu have some but not all of the features which make corona so transmissible – SARS-1 for example had no latent transmission while bird flu was extremely transmissible among birds (like unmasked people for corona) but less so among humans (like masked people). These more benign viruses can also be modelled easily using our approach. The public health interventions we have studied – masking, contact tracing and testing – are common to any infectious disease; they are about all we can throw at an emerging pathogen when there is no vaccine or cure. The immune response is also completely general – between the various possibilities, we have likely covered every possible infection, known or unknown. Of course, the model in its present form is not applicable to vector-borne diseases such as malaria and dengue. Scalar to vector extension in any physical problem is hasslesome but never undoable; the same should be true here.

Thus, although we have written this Article in terms of the specific disease COVID-19, we do not intend it to be a use-and-throw whose relevance will cease as soon as corona becomes over (though we have no idea at present as to when **that** happy day will arrive). Infectious disease has always been a part of human existence, and with the advent of jetliner travel, pathogens can be carried halfway across the world in a matter of hours. With current trends continuing, it is highly probable that pandemics are here to stay. We hope that the same may be said of our model as well.

## Data Availability

There is NO data referred to in the manuscript.

## Notes

### Competing Interest Statement

The authors have declared no competing interest.

### Clinical Trial

N/A

### Funding Statement

There is NO funding to report for this study.

### Author Declarations

No IRB approvals required due to mathematical modeling study

## REFERENCES

[1] https://cmmid.github.io/topics/covid19/

[2] https://www.imperial.ac.uk/mrc-global-infectious-disease-analysis/covid-19/covid-19-publications/

[3] https://covid-19.bsvgateway.org/

[4] S Thurner, P Klimer and R Hanel, “A Network-based explanation of why most COVID-19 infection curves are linear,” PNAS 117 (37), 22684-22689 (2020)

[5] JM Cashore et. al., “COVID-19 mathematical modeling for Cornell’s fall semester,” (2020) available at https://cpb-us-w2.wpmucdn.com/sites.coecis.cornell.edu/dist/3/341/files/2020/10/COVID_19_Modeling_Jun15-VD.pdf

[6] D Adak, A Majumder and N Bairagi, “Mathematical perspective of COVID-19 pandemic : disease extinction criteria in deterministic and stochastic models,” MedRxiv Article (2020) available at https://www.medrxiv.org/content/10.1101/2020.10.12.20211201v1

[7] Y Gu, “Estimating true infections : a simple heuristic to measure implied infection fatality rate,” (2020) available at https://covid19-projections.com/estimating-true-infections/

[8] WO Kermack and AG McKendrick, “A Contribution to the mathematical theory of epidemics,” Proceedings of the Royal Society A 115 (772), 700-721 (1927)

[9] G Giordano et. al., “Modeling the COVID-19 epidemic and implementations of population-wide interventions in Italy,” Nature Medicine 26 (6), 855-860 (2020)

[10] A Das, A Dhar, S Goyal and A Kundu, “COVID-19 : analysis of an extended SEIR model and comparison of different interventions strategies,” Arxiv Article 2005.11511 (2020)

[11] M Agrawal, M Kanitkar and M Vidyasagar, “Modeling the spread of SARS-CoV-2 pandemic : impact of lockdowns and interventions,” in press, Indian Journal of Medical Research (2020)

[12] G Menon, “Problems with the Indian supermodel for COVID-19,” available at https://www.thehindu.com/sci-tech/science/problems-with-the-indian-supermodel-for-covid-19/article32937184.ece

[13] R Singh and R Adhikari, “Age-structured impact of social distancing on the COVID-19 epidemic in India,” Arxiv Article 2003.12055 (2020)

[14] A Dhar, “A Critique of the COVID-19 analysis for India by Singh and Adhikari,” ibid. 2004.05373 (2020)

[15] LS Young, S Ruschel, S Yachuk and T Pereira, “Consequences of delays and imperfect implementation of isolation in epidemic control,” Scientific Reports 9, 3505 (2019)

[16] CP Vyasarayani and A Chatterjee, “New approximations and policy implications from a delayed dynamic model of a fast pandemic,” Physica D 414, 132701 (2020)

[17] L Dell’Anna, “Solvable delay model for epidemic spreading : the case of COVID-19 in Italy,” Arxiv Article 2003.13571 (2020)

[18] B Shayak, MM Sharma and M Gaur, “A New delay differential equation for COVID-19,” Proceedings of KIML Workshop, KDD2020

[19] MM Sharma and B Shayak, “Public health implications of a delay differential equation model for COVID-19,” ibid. (2020)

[20] B Shayak, MM Sharma, RH Rand, A Singh and A Misra, “A Delay differential equation model for the spread of COVID-19,” International Journal of Engineering Research and Applications 10 (10/3), 1–13 (2020)

[21] ML Childs et. al., “The Impact of long term non-pharmaceutical interventions on COVID-19 epidemic dynamics and control,” MedRxiv Article (2020) available at https://www.medrxiv.org/content/10.1101/2020.05.03.20089078v1

[22] B Shayak and RH Rand, “Self-burnout – a new path to the end of COVID-19,” ibid. (2020) available at https://www.medrxiv.org/content/10.1101/2020.04.17.20069443v2

[23] O Diekmann, JAP Heesterbeek and JAJ Metz, “On the Definition and computation of the basic reproduction ratio R0 in models for infectious diseases in heterogeneous populations,” Journal of Mathematical Biology 28 (4), 365–382 (1990)

[24] B Shayak, “Differential Equations – Linear Theory and Applications,” available electronically at www.shayak.in/Shayakpapers/DELTA/DELTA.pdf

[25] D Adjodah et. al., “Decrease in hospitalizations for COVID-19 after mask mandates in 1083 US counties,” MedRxiv Article (2020) available at https://www.medrxiv.org/content/10.1101/2020.10.21.20208728v1

[26] B Shayak, MM Sharma, RH Rand, A Singh and A Misra, “Transmission dynamics of COVID-19 and impact on public health policy,” ibid. (2020) available at https://www.medrxiv.org/content/10.1101/2020.03.29.20047035v1

[27] LM Kucirka, SA Lauer, O Laeyendecker, D Boon and J Lessler, “Variation in false-negative rate of RT-PCR tests for SARS-CoV02 by time since exposure,” ibid. (2020) available at https://www.medrxiv.org/content/10.1101/2020.04.07.20051474v1

[28] A Wajnberg et. al., “SARS-CoV-2 infection induces robust neutralizing antibody responses that are stable for at least three months,” ibid. (2020) available at https://www.medrxiv.org/content/10.1101/2020.07.14.20151126v1

[29] AWD Edridge et. al., “Human coronavirus reinfection dynamics : lessons for SARS-CoV-2,” ibid. (2020) available at https://www.medrxiv.org/content/10.1101/2020.05.11.20086439v2

[30] RJ Kosinski, “The Influence of time-limited immunity on a COVID-19 epidemic: a simulation study,” ibid. (2020) available at https://www.medrxiv.org/content/10.1101/2020.06.28.20142141v1

[31] F Sandmann et. al., “The Potential health and economic value of SARS-CoV-2 vaccination alongside physical distancing in the UK : transmission model-based future scenario analysis and economic evaluation,” ibid. (2020) available at https://www.medrxiv.org/content/10.1101/2020.09.24.20200857v1

[32] J Lavine, O Bjornstadt and R Antia, “Immunological characteristics will govern the changing severity of COVID-19 during the likely transition to endemicity,” ibid. (2020) available at https://www.medrxiv.org/content/10.1101/2020.09.03.20187856v1

[33] V Chin et. al., “A Case study in model failure – COVID-19 daily deaths and ICU bed utilization predictions in New York State,” Arxiv Article 2006.15997 (2020)

[34] SR Serrao et. al., “Requirements for the containment of COVID-19 disease outbreaks through periodic testing, isolation and quarantine,” MedRxiv Article (2020) available at https://www.medrxiv.org/content/10.1101/2020.10.21.20217331v1

